# Suicide deaths during the COVID-19 pandemic in the United States, by region, March 1, 2020-June 30, 2022

**DOI:** 10.1101/2023.01.18.23284681

**Authors:** Jeremy Samuel Faust, Benjamin Renton, Chengan Du, Sejal B. Shah, Alexander Junxiang Chen, Shu-Xia Li, Harlan M. Krumholz

## Abstract

**Introduction:** There were concerns that suicide deaths might increase due to Covid-19 pandemic-related stressors. Previous research demonstrated that suicide deaths actually decreased in 2020 in the US. An update covering 2021-2022 with regional data is warranted.

**Methods:** Observational cohort, US and regional data. Expected monthly deaths were modeled using pre-pandemic US and regional data (2015-2020). Mortality data was accessed from CDC public reporting.

**Results:** We find that suicide deaths in the United States were below expected levels throughout the pandemic period (March 1, 2020-June 30,2022) with >4,100 fewer suicide deaths than would have been expected to occur during the study period. Stratifying suicide mortality by US Census Bureau region yielded statistically significant decreases from expected suicide deaths in all regions except the Midwest, (which recorded no significant change in suicide deaths during the overall pandemic period).

**Conclusion:** Suicide mortality is down in the US since the pandemic began, through June 30, 2022. Possible explanations include an early “coming together” effect; Later, increased access to mental health resources and a greater focus on mental health in the media may have reduced stigma and barriers in seeking necessary psychiatric care.

## Introduction

Despite concerns that increased suicide deaths might occur due to isolation, mourning, and economic woes early in the COVID-19 pandemic, suicide rates in the US and other nations were lower than expected in 2020.^1,2^

However, the pandemic has lasted longer than anticipated. Longer-term stressors may have accumulated in areas where some social distancing continued, or, conversely, in areas which returned to normalcy but had higher death rates overall.

Accordingly, we studied suicide mortality during the COVID-19 pandemic in the US, by region, from March 1, 2020-June, 30, 2022.

## Methods

Using previously describing methods, we used seasonal autoregressive integrated moving average (sARIMA) models and applied them to population statistics (2014-2020), all-cause mortality, and suicide mortality data (January 2015-February 2020) to generate expected monthly populations and suicide deaths for March 1, 2020-June 30, 2022; populations were adjusted to reflect decreases due to cumulative all-cause mortality, as previously described. Excess suicide mortality was defined as the number of observed minus the number of expected deaths. 95% confidence intervals (CI) were calculated for each month and period. Data were acquired from CDC WONDER (finalized and provisional, ICD codes U03, X60–X84, Y87.0, underlying cause of death).^3^ We divided the pandemic into 9 periods: March 2020-June 2020 (4 months), and the 8 subsequent 3-month quarters for which there are public data. We studied the 4 Census regions and the entire US.

Analyses were conducted using R version 4.0.2. This study used publicly available data and was not subject to institutional review approval per HHS regulation 45 CFR 46.101(c).

## Results

During the pandemic, 115,082 suicide deaths were expected in the US; only 110,908 occurred (−4,174 excess deaths; 95% CI: −6387–−1962). Statistically significant decreases in suicide mortality occurred in 3 of 4 regions during the pandemic; the Northeast (^−^648; 95% CI: ^−^1267-^−^29), South (−1887; 95% CI: ^−^3096-^−^678), and West (^−^1537; 95% CI: ^−^2044-^−^1031), but not the Midwest (^−^102; 95% CI: ^−^570-366), (Figure). All regions had statistically significant decreases in suicide mortality during the pandemic portion of 2020; the West observed decreases in all pandemic years (Table).

Statistically significant decreases in suicide mortality occurred in 2 of 9 periods studied for the Northeast, Midwest, and South, and in 6 of 9 in the West (Table). Statistically significant increases occurred in 3 of 9 periods in the Midwest (Table). While the Midwest had significant increases during the first half of 2022, suicide deaths there remain statistically unchanged for the pandemic overall.

## Discussion

Despite early concerns, suicide deaths during the COVID-19 outbreak in the US (March 1, 2020-June 30, 2022) were statistically lower than projected, findings which were replicated in 3 of 4 census regions (the 4^th^ region recorded no overall change).

There are several possible explanations for these findings; a “coming together effect,” gave people a shared sense of purpose, early on. Later, mental health stressors may have mounted but increased care demands were counterbalanced by newly increased access to psychiatric resources previously unavailable, including telehealth and, in 2022, access to a universal national suicide hotline (988).^4^ Additionally, mental health has received more media attention recently, possibly decreasing stigma for seeking care, thereby leading to patient engagement with psychiatric resources, rather than suicidal behavior.^5^

This observational study has limitations. The findings cannot be explicitly explained by any particular characteristic of the COVID-19 pandemic. One confounding variable may be manner of death adjudication; some deaths assigned to unintentional overdose may have actually been suicides, and vice versa, as previously.^6^ However, baseline rates of misattribution are unlikely to have changed during the pandemic. Also, while the data are provisional, they are unlikely to change dramatically.

In sum, suicide deaths are lower in the US since the COVID-19 pandemic began. Maintaining expanded access to telehealth and other resources is paramount.

## Data Availability

All data produced in the present study are available upon reasonable request to the authors

## Author Contributions

Dr. Faust and Mr. Renton had full access to all of the data in the study and takes responsibility for the integrity of the data and the accuracy of the data analysis.

*Concept and design:* Faust, Du, Krumholz, Renton.

*Acquisition, analysis, or interpretation of data:* All authors.

*Drafting of the manuscript:* Renton, Shah, Faust.

*Critical revision of the manuscript for important intellectual content:* All authors.

*Statistical analysis:* Faust, Du, Li, Renton.

*Administrative, technical, or material support:* Chen, Renton

*Supervision:* Faust, Krumholz.

*Data visualization:* Renton.

## Conflict of Interest Disclosures

Dr. Krumholz reported receiving consulting fees from UnitedHealth, Element Science, Aetna, Reality Labs, F-Prime, and Tesseract/4Catalyst; serving as an expert witness for Martin/Baughman law firm, Arnold and Porter law firm, and Siegfried and Jensen law firm; being a cofounder of Hugo Health, a personal health information platform; being a cofounder of Refactor Health, an enterprise health care, artificial intelligence–augmented data management company; receiving contracts from the Centers for Medicare & Medicaid Services through Yale New Haven Hospital to develop and maintain performance measures that are publicly reported; and receiving grants from Johnson & Johnson outside the submitted work. No other disclosures were reported.

**Figure.**
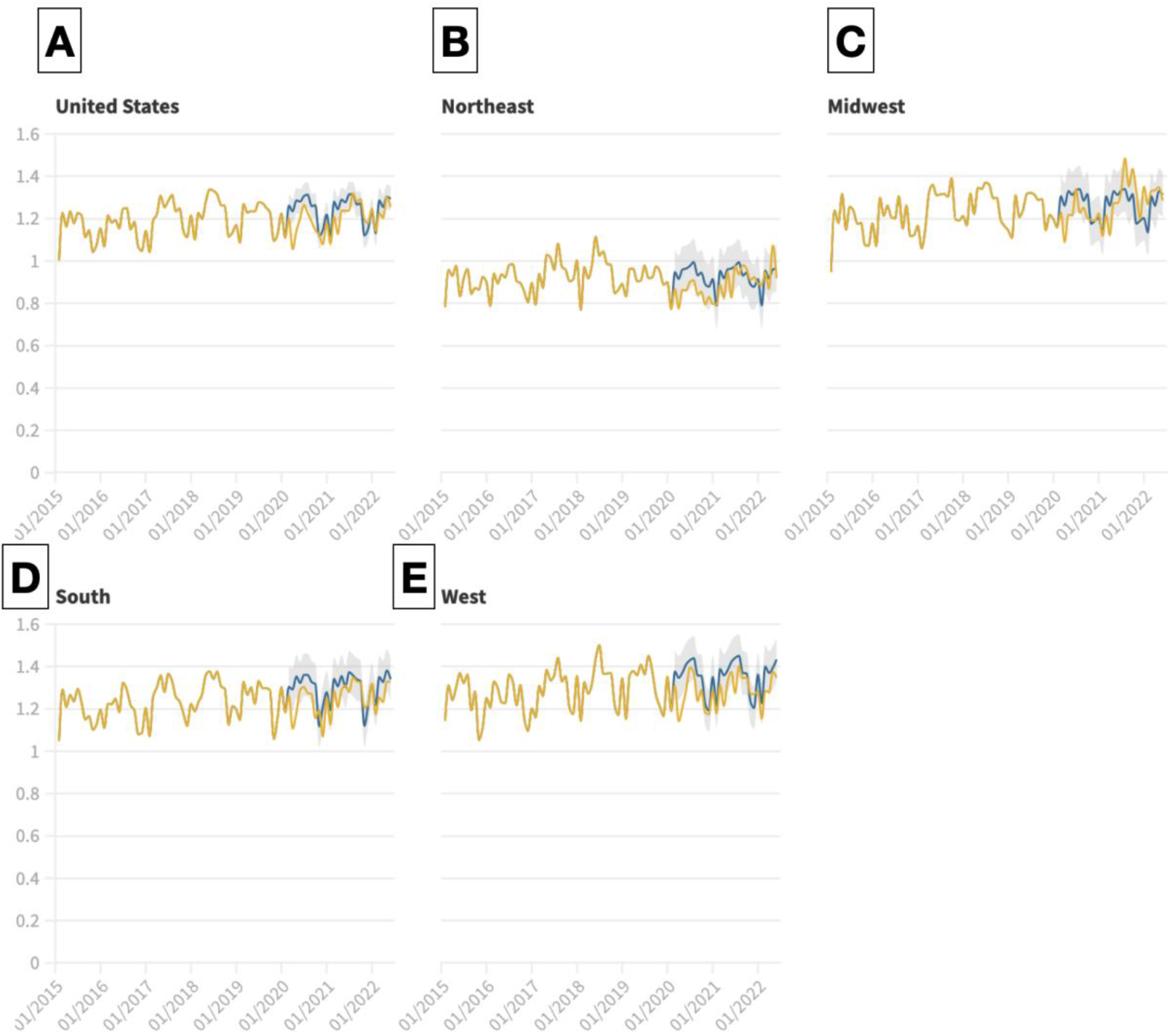
Incident rates of monthly observed suicide deaths per 100,000 population (yellow lines, January 1, 2015-June 30, 2022) and the number of expected deaths (blue lines, March 1, 2020-June 30, 2022) with 95% confidence intervals for expected deaths (gray shaded regions, March 1, 2020-June 30, 2022) generated using the seasonal adjusted model. For the United States (panel A), Northeast (panel B), Midwest (panel C), South (panel D), and West (panel E).

**Table.**
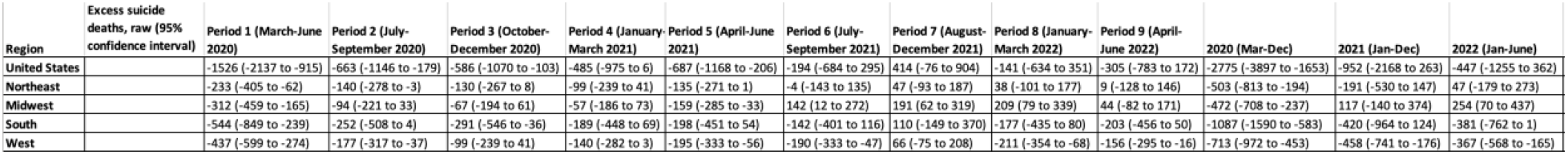

## Notes

### Funding Statement

This study did not receive any funding

### Author Declarations

This study used publicly available data from CDC WONDER and was not subject to institutional review approval per HHS regulation 45 CFR 46.101(c). https://wonder.cdc.gov/mcd-icd10-provisional.html

## References

1. Faust JS, D. C, Mayes KD, et al. Mortality From Drug Overdoses, Homicides, Unintentional Injuries, Motor Vehicle Crashes, and Suicides During the Pandemic, March-August 2020. JAMA. 2021;326(1):84–86. doi:10.1001/jama.2021.8012

2. Pirkis J, John A, Shin S, et al. Suicide trends in the early months of the COVID-19 pandemic: an interrupted time-series analysis of preliminary data from 21 countries. Lancet Psychiatry. 2021;8(7):579–588. doi:10.1016/S2215-0366(21)00091-2

3. National Center for Health Statistics. CDC WONDER. Accessed January 16, 2023. https://wonder.cdc.gov/

4. Lo J, Panchal N, Mar 15 BFMP, 2022. Telehealth Has Played an Outsized Role Meeting Mental Health Needs During the COVID-19 Pandemic. KFF. Published March 15, 2022. Accessed January 16, 2023. https://www.kff.org/coronavirus-covid-19/issue-brief/telehealth-has-played-an-outsized-role-meeting-mental-health-needs-during-the-covid-19-pandemic/

5. Beckjord, E. Contributor: COVID-19 and Stigma About Mental Health—A Pandemic Silver Lining? AJMC. Accessed January 16, 2023. https://www.ajmc.com/view/contributor-covid-19-and-stigma-about-mental-health-a-pandemic-silver-lining-

6. Bohnert ASB, Ilgen MA. Understanding Links among Opioid Use, Overdose, and Suicide. N Engl J Med. 2019;380(1):71–79. doi:10.1056/NEJMra1802148

